# External validation of the CAST and rCAST score in patients with out-of-hospital cardiac arrest who underwent ECPR: A secondary analysis of the SAVE-J II study

**DOI:** 10.1101/2023.05.17.23290147

**Authors:** Kayo Misumi, Yoshihiro Hagiwara, Takuya Kimura, Toru Hifumi, Akihiko Inoue, Tetsuya Sakamoto, Yasuhiro Kuroda, Takayuki Ogura, the SAVE-J II study group

## Abstract

**Background:** Risk stratification is important in patients with postcardiac arrest syndrome (PCAS). The post-Cardiac Arrest Syndrome for Therapeutic hypothermia (CAST) and revised CAST (rCAST) scores have been well validated for predicting the accuracy of neurological outcomes, particularly for conventionally resuscitated PCAS patients. However, no studies have evaluated patients undergoing extracorporeal cardiopulmonary resuscitation (ECPR).

**Methods:** Adult patients with out-of-hospital cardiac arrest (OHCA) who underwent ECPR were analyzed in this retrospective observational multicenter cohort study (SAVE-J II study). We validated the accuracy of the CAST and rCAST scores for predicting favorable neurological outcomes (cerebral performance category 1 or 2) at 30 days. Moreover, we compared the predictive performance of these CAST/rCAST scores with those of the previously documented TiPS65 risk model derived from patients with OHCA who were resuscitated using ECPR.

**Results:** A total of 1135 patients were analyzed. Their median age was 60 years and males comprised 84% of the cohort. The proportion of patients with favorable neurological outcomes was 16.6% overall. In the external validation, the area under the curve (AUC) of the CAST score was numerically larger than those of the rCAST and TiPS65 scores (AUC 0.677 vs. 0.603: p < 0.001, vs. 0.633: p = 0.154, respectively). Both CAST/rCAST risk scores showed good calibration (Hosmer– Lemeshow test: p = 0.726 and 0.674, respectively). Furthermore, the CAST score showed significantly better predictability in net reclassification than did the rCAST (p < 0.001) and TiPS65 scores (p = 0.001).

**Conclusions:** CAST and rCAST scores can predict neurological outcomes in patients with OHCA who undergo ECPR. The prognostic accuracy of the CAST score was significantly better than that of other pre-existing risk prediction models.

**IRB information:** The present study was approved by the institutional review board of Kagawa University (approval number: 2018-110)

**Clinical trial registration:** https://center6.umin.ac.jp/cgi-open-bin/ctr_e/ctr_view.cgi?recptno=R000041577 (unique identifier: UMIN000036490)

## Introduction

Although cardiovascular disease prevention and prehospital medical systems have been developed in recent decades, the number of patients with out-of-hospital cardiac arrest (OHCA) remains high[1-3], and the related medical expenses have also increased[3,4]. The severity of postcardiac arrest syndrome (PCAS) varies with the duration and cause of cardiac arrest. Accurate prediction of neurological outcomes in patients with PCAS is frequently uncertain; however, it is one of the most important factors in the proper allocation of medical resources and in avoiding premature withdrawal of life-sustaining therapies in patients who may yet achieve a good neurological outcome.

Extracorporeal cardiopulmonary resuscitation (ECPR) is a method that uses extracorporeal membrane oxygenation (ECMO) for resuscitating patients who have not achieved return of spontaneous circulation (ROSC) by conventional cardiopulmonary resuscitation[5]. It has recently been suggested that ECPR can be useful for patients with cardiac arrest[6,7], and the number of patients with OHCA who undergo ECPR is increasing.

To recognize the severity of neurological damage at the time of ICU admission, the post-Cardiac Arrest Syndrome for induced Therapeutic hypothermia (CAST) score and revised CAST (rCAST) score have been proposed to predict neurological outcomes in patients with PCAS [8-10]. These scores were developed in adult patient cohorts, and has been well validated in patients with several different backgrounds[10-12]. However, the CAST and rCAST risk models have never been validated in patients with OHCA who underwent ECPR.

Therefore, this study aimed to evaluate the accuracy of CAST and rCAST scores in predicting neurological outcomes in patients who have undergone ECPR, and to compare the performance of these scores with those of other pre-existing risk models.

## Methods

### Study design and cohort

To examine whether the preexisting risk scores were adaptable to patients with OHCA who underwent ECPR, we used a data set from the SAJV-J II study cohort. The SAJV-J II study design has been reported elsewhere in detail[13,14]. In brief, this registry was a retrospective multicenter study conducted in Japan (involving 36 university and community hospitals), enrolling 2,157 consecutive patients with OHCA aged ≥ 18 years who were resuscitated using ECPR between January 1, 2013, and December 31, 2018. In this study, ECPR was defined as resuscitation with veno-arterial extracorporeal membrane oxygenation (VA-ECMO) in patients with refractory cardiac arrest.

This study was approved by the Kagawa University Institutional Ethics Committee (approval number: 2018-110) and by that of each participating institution. This secondary analysis of de-identified data was approved by the Institutional Review Board of the Saiseikai Utsunomiya Hospital (approval number: 2023-01). The requirement for informed consent was waived by the ethics committee because of the retrospective nature of the study. This study was performed in accordance with the 1975 Declaration of Helsinki Guidelines for Clinical Research Protocols.

### Study population

In this study, all patients in the SAVE-J II registry were included. The exclusion criteria were as follows: implementation of ECPR after intensive care unit admission; non-cardiac conditions, including acute aortic syndrome/aortic aneurysm, hypothermia, primary cerebral disorder, infection, drug intoxication, trauma, suffocation, drowning, and other external causes; patients who achieved ROSC at hospital arrival patients or who achieved ROSC before cannulation; OHCA patients who were transferred to the participating institutions from another hospital; those who withdrew after cannulation and before turning the ECMO pump on due to the return of spontaneous circulation (ROSC); patients with missing outcome data[13]; and patients with missing data on any of components required to calculate the CAST or rCAST scores and TiPS65 score.

### Outcome measurements

The primary endpoint of this study was favorable neurological outcomes at 30 days. We used the Cerebral Performance Categories (CPC) scale at 30 days as the primary outcome, as follows: CPC 1, full recovery; CPC 2, moderate disability; CPC 3, severe disability; CPC 4, coma or vegetative state; and CPC 5, death. CPC 1–2 was considered a favorable outcome, and CPC 3–5 represented an unfavorable outcome[15].

### Calculation of CAST score, rCAST score and TiPS65 score

In this study, we used CAST and rCAST scores to predict neurological outcomes. Both scores were calculated using variables obtained at the time of hospital arrival and ROSC. Moreover, we used the TiPS65 score as a comparator for these two risk scores. The TiPS65 score was derived from and validated in patients treated with ECPR[16,17]. The formulae used to calculate each risk score are shown in **Supplemental Figure S1**. In short, the CAST score was calculated from eight values: initial rhythm, presence/absence of witness/time until ROSC, blood pH upon hospital arrival, blood lactate level upon hospital arrival, Glasgow Coma Scale (GCS) motor score at the time of ROSC, serum albumin level, serum hemoglobin level, and gray matter:white matter ratio (GWR) [8]. The rCAST score, which is a simplified version of the original CAST score, comprises only the first five variables mentioned above and simplified coefficients[10]. The TiPS65 score is composed of four variables (Ti, time from the call for an ambulance to hospital arrival, ≤ 25 min; P, pH value on admission, ≥ 7.0; S, shockable rhythm on hospital arrival; and 65, age ≤ 65 years.10). One point was assigned to each of these four predictors, with the total score ranging from 0 to 4 points[16].

### Statistical analysis

Continuous variables are expressed as medians with interquartile ranges. Categorical variables are expressed as numbers and percentages. For validation, we calculated CAST and rCAST scores to evaluate the accuracy of the models. The discrimination of each risk score was verified by plotting the receiver operating characteristic (ROC) curve and evaluating the area under the ROC curve (AUC). We calculated the observed outcome probabilities with 95% confidence intervals (CIs). In addition, to check whether the performance of the CAST score was adaptable to ECPR patients, we visually compared the observed outcomes and used a bar plot in which patients were divided into four quartile groups. For calibration, the Hosmer–Lemeshow test was performed.

Moreover, to compare the performance of these two CAST/rCAST risk models with the preexisting TiPS65 risk model, discrimination and net reclassification improvements were calculated among these three risk scores[18].

Differences were considered statistically significant at two-tailed P-values < 0.05. All analyses were performed using R software (version 4.2.1; R Foundation for Statistical Computing, Vienna, Austria).

## Results

Among the 2,157 patients registered in the SAJV-J II study, 1,135 were included and analyzed (**Figure 1**). The baseline patient characteristics of all cohorts and the comparison between CPC 1–2 and CPC 3–5 are shown in **Table 1**. The median age of patients was 60 years. Overall, 951 (83.8%) patients were men. The proportion of patients with favorable neurological outcomes at 30 days was 16.6%. The survival rate at 30 days was 33.7% overall. The group with favorable neurological outcomes had a higher proportion of patients receiving bystander CPR and with an initial shockable rhythm.

**Table 1.** Patient characteristics

| Variables | Overall, (n =1,135) | 30-day neurological outcome |  |
| --- | --- | --- | --- |
|  |  | Favorable, (n=188) | Unfavorable, (n=947) |
| Age, years | 60 (49 ,69) | 55 (44, 66) | 61 (50, 69) |
| Sex Male, n (%) | 951 (84 %) | 144 (77 %) | 807 (85 %) |
| Witness, n (%) | 897 (79 %) | 158 (84 %) | 739 (78 %) |
| Bystander CPR, n (%) | 651/1128 (57 %) | 129/188 (69 %) | 522/940 (56 %) |
| Initial rhythm, shockable, n (%) | 794 (70 %) | 152 (81 %) | 642 (68 %) |
| Estimated low flow time (n=1092) | 53 (43, 63) | 50 (40, 62) | 53 (44, 64) |
| Time from onset to ECMO (n=1001) | 56 (46, 69) | 55 (42, 66) | 56 (47, 69) |
| **Cause of cardiac arrest** |  |  |  |
| Cardiac | 983 (87 %) | 163 (87 %) | 820 (87 %) |
| Non-cardiac internal | 84 (7.4 %) | 17 (9.0 %) | 67 (7.1 %) |
| Internal-unknown | 13 (1.2 %) | 5 (2.7 %) | 8 (0.8 %) |
| Unknown | 55 (4.9 %) | 3 (1.6 %) | 52 (5.5 %) |
| **Data at the time of ROSC** |  |  |  |
| GCS M, $\geq 2$, n (%) | 11 (1.0 %) | 8 (4.3 %) | 3 (0.3 %) |
| Serum pH | 6.93 (6.81, 7.04) | 6.96 (6.34, 7.10) | 6.93 (6.81, 7.03) |
| Serum lactate, mmol/dL | 13.0 (10.2, 16.0) | 12.8 (9.7, 16.0) | 13.1 (10.2, 16.0) |
| Albumin, g/dL | 3.1 (2.6, 3.5) | 3.3 (1.2, 3.8) | 3.0 (2.6, 3.5) |
| Hemoglobin, g/dL | 12.7 (11.0, 14.5) | 13.3 (11.7, 15.3) | 12.6 (10.9, 14.4) |
| **Targeted Temperature Management** |  |  |  |
| Hypothermia (TTM $< 35^{\circ}\text{C}$) | 595 (52 %) | 113 (60 %) | 482 (51%) |
| Normothermia (TTM at $35\text{-}37^{\circ}\text{C}$) | 276 (24 %) | 50 (27 %) | 226 (24 %) |
| Unknown | 264 (23 %) | 25 (13 %) | 239 (25 %) |
| Survival at 30 days, n (%) | 383 (34 %) | 188 (100 %) | 195 (20.6 %) |

Data are presented as the median [interquartile range], or n (%). CPR, Cardiopulmonary resuscitation; ECMO, extracorporeal membrane oxygenation; ROSC, resumption of spontaneous heartbeat; GCS, Glasgow Coma Scale; TTM, Targeted Temperature Management.

**Figure 1.**
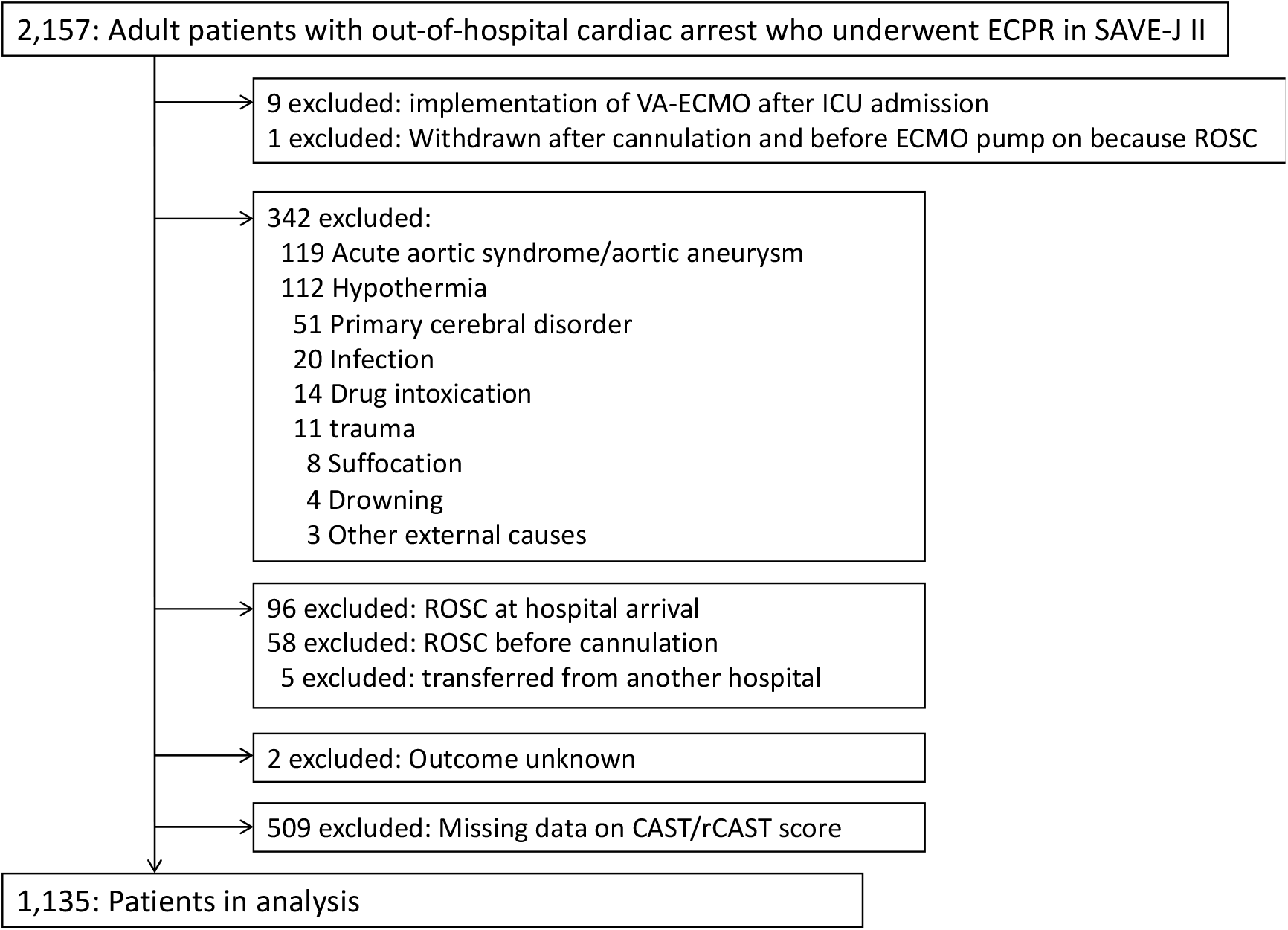
Flowchart of enrollment of study participants. ECPR, extracorporeal cardiopulmonary resuscitation; ECMO, extracorporeal membrane oxygenation; ICU, intensive care unit; ROSC, return of spontaneous circulation; CAST, post-Cardiac Arrest Syndrome for Therapeutic hypothermia; rCAST, revised CAST.

In the external validation, the areas under the curve (AUC) of the CAST and rCAST scores were 0.677 and 0.603, respectively (**Figure 2**). The calibration test was performed by applying the CAST and rCAST scores, which showed that both scores showed good calibration (Hosmer–Lemeshow test: p = 0.726 and 0.674, respectively).

**Figure 2.**
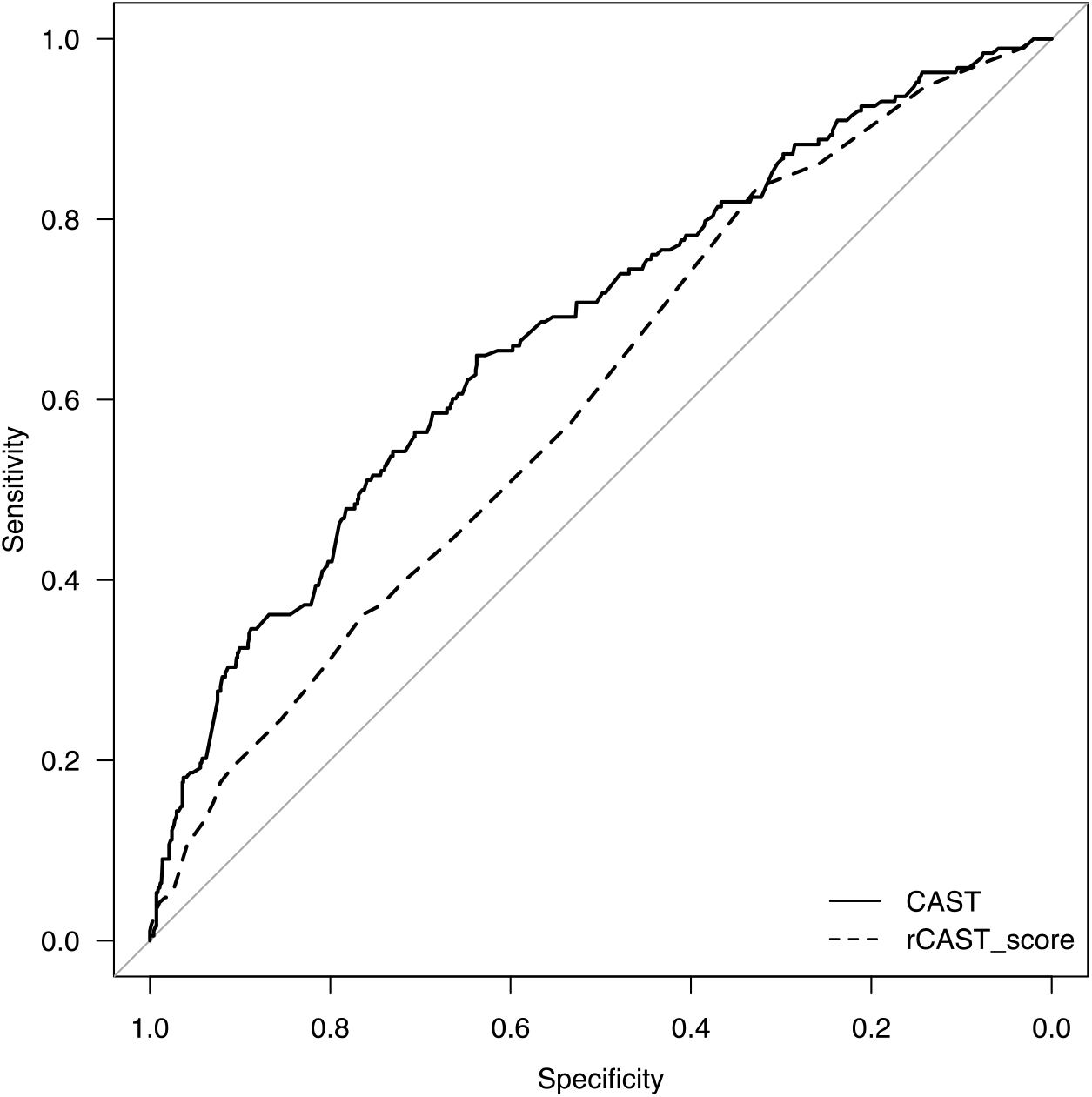
Receiver operator characteristic curves for the CAST and rCAST scores. CAST, post-Cardiac Arrest Syndrome for Therapeutic hypothermia; rCAST, revised CAST.

To check whether the performance of the CAST score was consistent in patients receiving ECPR, the total cohort was divided into four groups according to the CAST score quartile. The proportions of observed outcomes with 95% CIs for CAST scores in each group were 29.4% (24.1–35.2%), 17.1% (13.0–21.9%), 12.9% (9.2–17.4%), and 7.5% (4.8–11.2%) (**Figure 3**).

**Figure 3.**
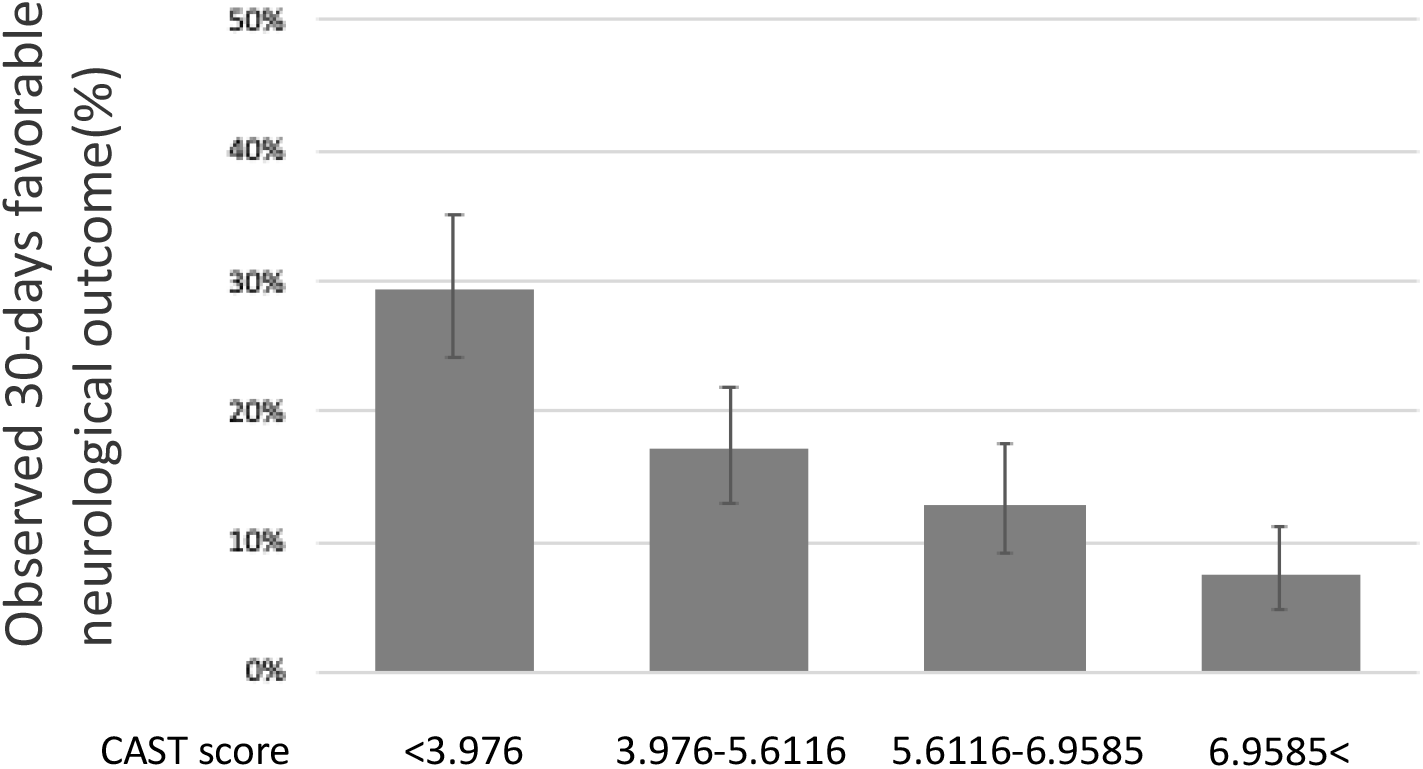
The proportion of observed favorable neurological outcome at 30 days in SAVE-J II study stratified by risk groups in CAST score. CAST, post-Cardiac Arrest Syndrome for Therapeutic hypothermia.

Finally, we compared the CAST, rCAST, and TiPS65 scores to clarify the predictability of neurological outcomes. Since some variables of the TiPS65 score were not available for some patients (missing data: n = 12), we analyzed 1,123 patients and compared the TiPS65 score with the CAST and rCAST scores obtained from the same patients, completely excluding those with missing data. The CAST score was significantly better than the rCAST score in terms of discrimination (0.677 vs. 0.603, p < 0.001) and reclassification (p < 0.001). Moreover, although no statistically significant difference in AUCs was observed between the CATS and TiPS65 scores, the CAST score showed a significant net improvement in reclassification (p = 0.001, **Table 2**).

**Table 2.**
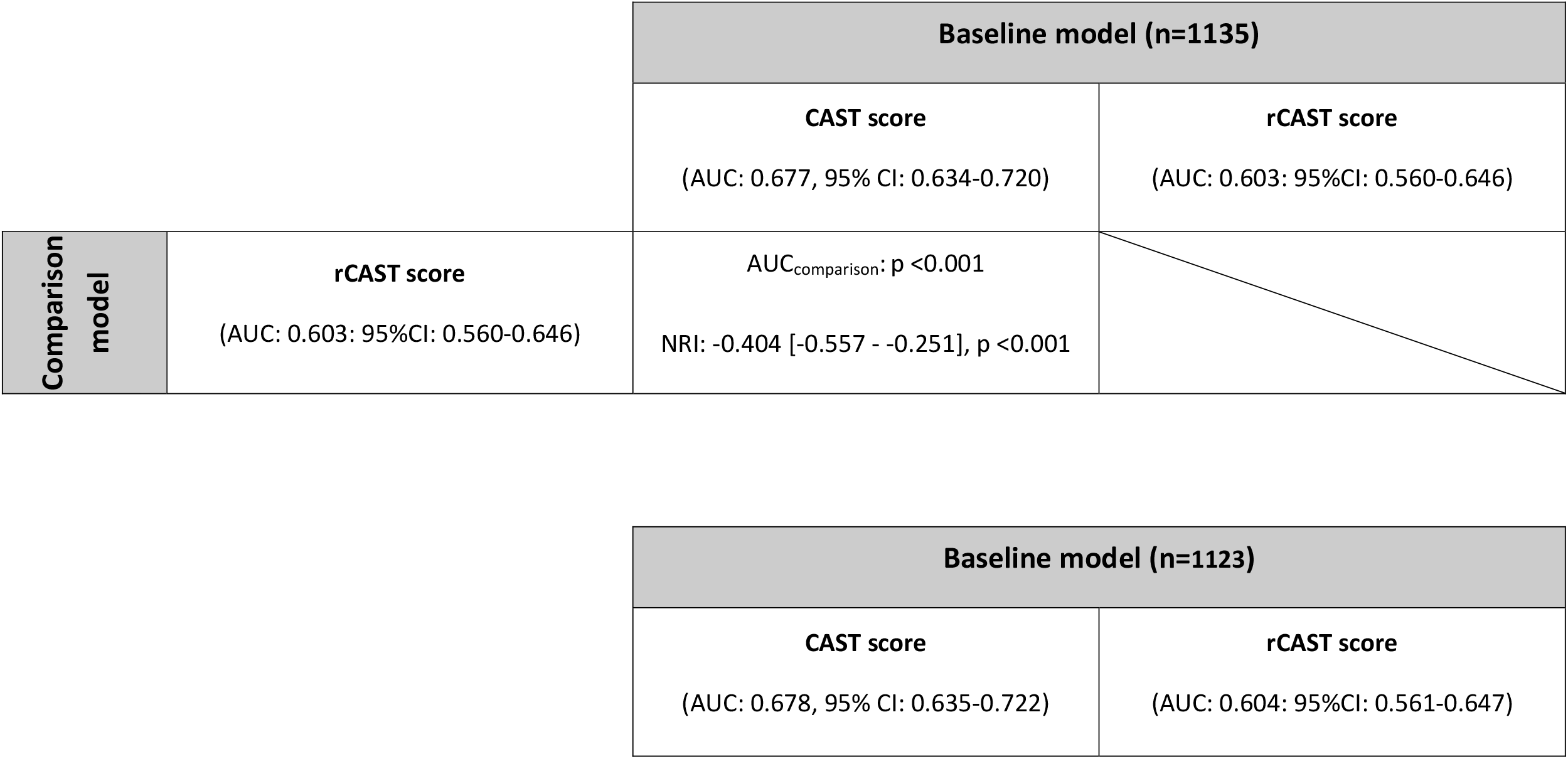
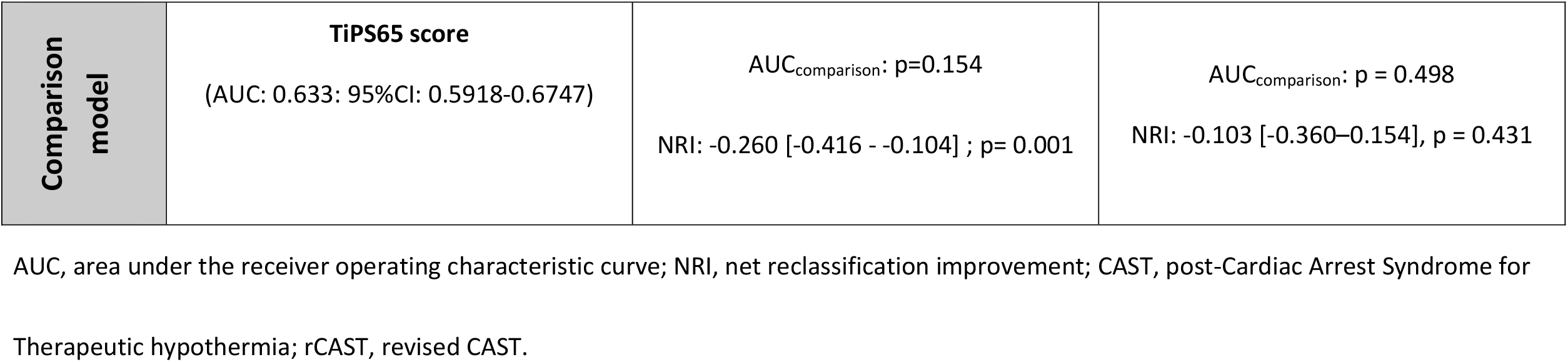
Comparison of the AUCs and net reclassification improvement of the CAST, rCAST, and TiPS65 risk models for neurological outcome at 30 days.

## Discussion

In this study, we tested the predictive ability of the CAST and rCAST scores for estimating neurological outcomes at 30 days in patients with PCAS resuscitated using ECPR. These two risk models showed moderate discrimination and good calibration in the largest OHCA ECPR registry in Japan, SAVE-J II. Furthermore, we compared the performances of these risk models with each other and with the TiPS65 score. The CAST score performed better than did the other two risk models.

The CAST and rCAST risk models were originally developed and validated in cohorts composed of adult patients who had been resuscitated with conventional cardiopulmonary resuscitation, that is, without ECPR, after experiencing cardiac arrest[8,10]. ECPR is an approach for resuscitation using ECMO in patients who do not achieve ROSC by conventional cardiopulmonary resuscitation[5]. In these patients, the severity of their condition tends to be higher due to the longer duration of the resuscitation, and the backgrounds of patients who underwent ECRP were different from those who did not, particularly in a retrospective cohort[19,20]. Previous studies showed that the AUC of the CAST score was 0.97[8] and that of the rCAST score was around 0.8–0.9 [10-12] for predicting neurological outcomes at 30 days. The present study showed that the AUC of the CAST score was 0.677 in the patients who underwent ECPR, which was lower than that reported in previous studies, but still indicated a moderate predictive value.

Moreover, the TiPS65 risk model was originally derived from patients undergoing ECPR, and its prediction accuracy was moderate. The TiPS65 score is user-friendly and consists of only four integer parameters. However, the prognostic value of the CAST score was numerically superior to that of the TiPS65. Although the formula for calculating the CAST score is complicated, tools for calculating the CAST score are available as an iOS application: iPad: https://geo.itunes.apple.com/jp/app/meidaiscore-for-ipad/id1065338535?mt = 8; iPhone: https://geo.itunes.apple.com/jp/app/meidai-score-for-iphone/id1067612773?mt = 8. This may help physicians and intensivists to calculate the prognosis of the patient prior to ECPR administration in an emergency room.

In this study, it was not considered whether patients in the cohort had been undergoing targeted temperature management (TTM). Although this may possible influence the neurological outcome, the proportion of patients undergoing TTM and the target temperature did not differ markedly between the favorable and unfavorable outcome groups. Moreover, earlier randomized controlled trials have reported no significant difference in neurological outcomes between hypothermic and normothermic management[21,22]. Under ECMO management in this study, almost all patients were expected to have avoided hyperthermia due to the heat exchanger of the ECMO.

### Limitation

This study had several limitations. First, as this was a retrospective observational study; the inclusion criteria were not defined, and the indications for performing ECPR, such as patient selection and timing of ECMO insertion, were dependent on each physiatrist and institution. Second, we defined low-flow time as the duration from the beginning of CPR to the initiation of the ECMO pump. However, this definition is different from that of the actual low-flow time. Third, we used a Japanese cohort, and the Japanese emergency medical system is somewhat different from American and European countries in terms of the point transfer protocol, including the timing of drug administration. Thus, the applicability of our findings to other nationalities remains unclear. Hence, further investigations with prospective large-population cohorts of various nationalities are required.

## Conclusion

We showed that the CAST and rCAST scores might have moderate predictive value for indicating favorable neurological prognosis in patients with PCAS who are resuscitated using ECPR. The CAST risk model showed better discrimination ability in terms of predicting outcomes than did the rCAST and TiPS65 risk models.

## Data Availability

Raw data were generated at Saiseikai Utsunomiya Hospital. Derived data supporting the findings of this study are available from the corresponding author KM on request.

## Acknowledgments

The authors would like to thank all members of the SAVE-J II study group who participated in this study: Hirotaka Sawano, M.D., Ph.D. (Osaka Saiseikai Senri Hospital), Yuko Egawa, M.D., Shunichi Kato, M.D. (Saitama Red Cross Hospital), Kazuhiro Sugiyama M.D. (Tokyo Metropolitan Bokutoh Hospital), Naofumi Bunya, M.D., Takehiko Kasai, M.D. (Sapporo Medical University), Shinichi Ijuin, M.D., Shinichi Nakayama, M.D., Ph.D. (Hyogo Emergency Medical Center), Jun Kanda, M.D., Ph.D., Seiya Kanou, M.D. (Teikyo University Hospital), Toru Takiguchi, M.D., Shoji Yokobori, M.D., Ph.D. (Nippon Medical School), Hiroaki Takada, M.D., Kazushige Inoue, M.D. (National Hospital Organization Disaster Medical Center), Ichiro Takeuchi, M.D., Ph.D., Hiroshi Honzawa, M.D. (Yokohama City University Medical Center), Makoto Kobayashi, M.D., Ph.D., Tomohiro Hamagami, M.D. (Toyooka Public Hospital), Wataru Takayama, M.D., Yasuhiro Otomo, M.D., Ph.D. (Tokyo Medical and Dental University Hospital of Medicine), Kunihiko Maekawa, M.D. (Hokkaido University Hospital), Takafumi Shimizu, M.D., Satoshi Nara, M.D. (Teine Keijinkai Hospital), Michitaka Nasu, M.D., Kuniko Takahashi, M.D. (Urasoe General Hospital), Yoshihiro Hagiwara, M.D., M.P.H. (Imperial Foundation Saiseikai, Utsunomiya Hospital), Reo Fukuda, M.D. (Nippon Medical School Tama Nagayama Hospital), Takayuki Ogura, M.D., Ph.D. (Japan Red Cross Maebashi Hospital), Shinichiro Shiraishi, M.D. (Aizu Central Hospital), Ryosuke Zushi, M.D. (Osaka Mishima Emergency Critical Care Center), Norio Otani, M.D. (St. Luke’s International Hospital), Migaku Kikuchi, M.D., Ph.D. (Dokkyo Medical University), Kazuhiro Watanabe, M.D. (Nihon University Hospital), Takuo Nakagami, M.D. (Omihachiman Community Medical Center), Tomohisa Shoko, M.D., Ph.D. (Tokyo Women’s Medical University Medical Center East), Nobuya Kitamura, M.D., Ph.D. (Kimitsu Chuo Hospital), Takayuki Otani, M.D. (Hiroshima City Hiroshima Citizens Hospital), Yoshinori Matsuoka, M.D., Ph.D. (Kobe City Medical Center General Hospital), Masaaki Sakuraya, M.D., M.P.H. (JA Hiroshima General Hospital Hiroshima), Hideki Arimoto, M.D. (Osaka City General Hospital), Koichiro Homma, M.D., Ph.D. (Keio University School of Medicine), Hiromichi Naito, M.D., Ph.D. (Okayama University Hospital), Shunichiro Nakao, M.D., Ph.D. (Osaka University Graduate School of Medicine), Tomoya Okazaki, M.D., Ph.D. (Kagawa University Hospital), Yoshio Tahara, M.D., Ph.D. (National Cerebral and Cardiovascular Center), Hiroshi Okamoto, M.D, M.P.H. (St. Luke’s International Hospital), Jun Kunikata, M.D., Ph.D., and Hideto Yokoi, M.D., Ph.D. (Kagawa University Hospital).

## Material

Figure S1.

